# Lower regional gray matter volume in the absence of higher cortical amyloid burden in late-life depression

**DOI:** 10.1101/2021.02.08.21250568

**Authors:** Akihiro Takamiya, Thomas Vande Casteele, Michel Koole, François-Laurent De Winter, Filip Bouckaert, Jan Van den Stock, Stefan Sunaert, Patrick Dupont, Rik Vandenberghe, Koen Van Laere, Mathieu Vandenbulcke, Louise Emsell

## Abstract

Late-life depression (LLD) is associated with a risk of developing Alzheimer’s disease (AD). However, the role of AD-pathophysiology in LLD, and its association with clinical symptoms and cognitive function are elusive. In this study, one hundred subjects underwent amyloid positron emission tomography (PET) imaging with [^18^F]-flutemetamol and structural MRI: 48 severely depressed elderly subjects (age 74.1±7.5 years, 33 female) and 52 age-/gender-matched healthy controls (72.4±6.4 years, 37 female). The Geriatric Depression Scale (GDS) and Rey Auditory Verbal Learning Test (RAVLT) were used to assess the severity of depressive symptoms and episodic memory function respectively. Amyloid deposition was quantified using the standardized uptake value ratio. Whole-brain voxel-wise comparisons of amyloid deposition and gray matter volume (GMV) between LLD and controls were performed. Multivariate analysis of covariance was conducted to investigate the association of regional differences in amyloid deposition and GMV with clinical factors, including GDS and RAVLT. As a result, there were no significant group differences in amyloid deposition. In contrast, LLD showed significant lower GMV in the left temporal and parietal region. GMV reduction in the left temporal region was associated with episodic memory dysfunction, but not with depression severity. Regional GMV reduction was not associated with amyloid deposition. LLD is associated with lower GMV in regions that overlap with AD-pathophysiology, and which are associated with episodic memory function. The lack of corresponding associations with amyloid suggests that lower GM driven by non-amyloid pathology may play a central role in the neurobiology of LLD presenting as a psychiatric disorder.

## Introduction

Late-life depression (LLD) is a common but disabling psychiatric disorder with a high risk of mortality [1]. Given the global aging population, LLD has become a significant public health concern. LLD is associated with cognitive dysfunction, including episodic memory impairment [2–4], which may persist even after the depressive episode has been treated successfully [5, 6]. This and multiple lines of evidence associating LLD with an increased risk of Alzheimer’s disease (AD) [7, 8] suggest shared neuropathophysiology between the two disorders. The main neurobiological markers in this context are increased cerebral amyloid deposition, tau deposition, and gray matter (GM) atrophy, particularly in the medial temporal lobe (MTL).

Previous research into the relationship between amyloid deposition and depression has yielded mixed results. For example, neuropathological studies have reported both significant associations of amyloid deposition with depressive symptoms or a history of depression in patients with dementia [9, 10], and no associations [11, 12]. Longitudinal positron emission tomography (PET) imaging studies in cognitively normal subjects have consistently shown baseline amyloid burden to be associated with emerging mild depressive symptoms during follow-up [13–15], whereas others did not find any associations [16–18]. Notably, a recent large multisite florbetapir-PET study from the ADNI consortium reported lower, not higher, amyloid accumulation in LLD with mild depressive symptoms [19].

Both LLD and AD are associated with lower gray matter volume (GMV). In AD, GM atrophy is widespread with the most pronounced GM loss in the temporal and parietal cortex [20, 21]. Findings in LLD are inconsistent: lower GMV was reported in temporal [22, 23], prefrontal [22, 24], and parietal [23] regions, and in the hippocampus [24, 25], thalamus, and putamen [24], whereas some studies did not find any significant GMV reduction [26, 27]. This inconsistency might be due to the heterogeneity of LLD. For example, late-onset depression (LOD) may be related to vascular or neurodegenerative changes [28], whereas early-onset depression (EOD) may be more familial with a genetic predisposition [29]. In addition, psychotic depression, which is common in LLD [30], may be considered a distinct entity from non-psychotic depression [31].

Studies that have assessed the relationship between GMV and amyloid deposition in LLD are scarce. In a previous study, we reported lower hippocampal volume in LLD and found no association with amyloid deposition in regions associated with AD nor with episodic memory scores [32].

In sum, most neuropathological and neuroimaging studies so far have focused on the relationship between amyloid deposition and depressive symptoms or a history of depression in cognitively normal subjects and subjects who had already been diagnosed with MCI or AD. Although results from these studies provide some insight into the relationship between amyloid burden and depressive symptoms or with previous depressive episodes, they are less informative regarding the neurobiology of LLD as a psychiatric disorder. The distinction here is important, as depressive symptoms in healthy subjects or subjects with neurodegenerative disorders may have a different underlying neural substrate to the mood symptoms that present alongside the more severe and wide-ranging neuropsychological and psychomotor deficits that characterize LLD.

In this study, we extend our previous work by applying whole-brain voxel-wise analyses to identify regional differences in amyloid deposition and GMV between patients with LLD and healthy subjects. Additionally, we investigate how these regional differences relate to clinical factors, including episodic memory scores, the age at onset of depression, and the presence of psychosis.

## Materials and Methods

### Participants

This study was approved by the ethical committee of the University Hospitals Leuven (protocol S52151) and registered as a clinical trial (EudraCT Number 2009-018064-95). All participants provided written informed consent prior to enrollment in the study in accordance with the Declaration of Helsinki. Participants over 55 years of age with a diagnosis of major depressive disorder (MDD) according to the Diagnostic and Statistical Manual of Mental Disorders, Fourth Edition, Text Revision (DSM-IV-TR) were recruited at the University Psychiatric Center KU Leuven, Belgium. The exclusion criteria were: primary referral for assessment of cognitive impairment; comorbid major psychiatric illness; previous or current alcohol or drug dependence; neurological illness, including dementia; illness or medication precluding cognitive testing; metal implants precluding MRI scanning and ECT in the past 6 months. Fifty-two age- and gender-matched healthy comparison subjects were included, of which 46 were recruited from another [^18^F]-flutemetamol study at our center [33]. At inclusion, these subjects had a Clinical Dementia Rating score of 0 and Mini-Mental State Examination score >26 and were stratified for ApoE *ε*4 allele. By design almost half of this cohort (44.6%) was an ApoE *ε*4 carrier [33]. However, the ApoE *ε*4 status of the stratified comparison subjects did not differ from those of the patient group or the non-stratified comparison group, nor was there a difference in the proportion of amyloid positive participants defined by a standardized uptake value ratio (SUVR) threshold >1.38 (10 patients with LLD and 12 controls) [32]. In addition, the proportion of the ApoE *ε*4 carrier was not different between LOD (21.4%) and EOD (35.0%) (p = 0.34), nor between psychotic depression (37.5%) and non-psychotic depression (21.9%) (p = 0.31). The cohort of this study is the same as the previous study [32].

### Clinical assessment

The Geriatric Depression Scale (GDS) and Rey Auditory Verbal Learning Test (RAVLT) were used to assess the severity of depressive symptoms and episodic memory, respectively. Raw scores of neuropsychological data were converted to standardized z scores based on the mean and standard deviation of the healthy control group. The episodic memory domain score was calculated by summing the z-cores of total recall score (the total number of correctly recalled during the five trials), delayed recall score, and recognition score (the number of correct hits minus the number of false positives) [34]. This domain score was used to investigate the association between episodic memory and the imaging measures.

### MRI acquisition and image processing

High-resolution 3D turbo field echo (3DTFE) T1-weighted images were acquired using a 3-T Philips Intera scanner with an 8-channel head coil (Best, the Netherlands) with parameters: TR = 9.6s, TE = 4.6ms, flip angle = 8 degrees, Slice thickness = 1.2 mm, in-plane voxel-size = 0.98 × 0.98 × 1.2 mm^3^, 182 axial slices. Due to decommissioning of the 3T Intera scanner, 19 controls were scanned with identical sequence parameters on a 3T Philips Achieva scanner using a 32-channel head coil. To account for the effect of the different scanner on the analysis, we included scanner type as a covariate in the analysis, and we reanalyzed data after excluding these 19 subjects’ data.

All T1-images were processed using the default pipeline of the Computational Anatomy Toolbox (CAT12, http://dbm.neuro.uni-jena.de/cat/), a toolbox for Statistical Parametric Mapping software (SPM12, http://www.fil.ion.ucl.ac.uk/spm). Prior to preprocessing, all data were manually aligned with the anterior commissure–posterior commissure (AC-PC) line. Preprocessing included bias-correction, segmentation into GM, white matter (WM), and cerebrospinal fluid (CSF), spatial normalization using the DARTEL algorithm, and modulation. A study-specific DARTEL template was created from all participants’ data in this study. The pre-processed images were smoothed with a Gaussian kernel of 8-mm full-width at half maximum (FWHM). Total intracranial volume (TIV) was calculated using CAT12.

### Amyloid imaging

[^18^F]-Flutemetamol PET imaging was acquired on a 16-slice Siemens Biograph PET/CT scanner (Siemens, Erlangen, Germany) at the University Hospitals Leuven, as described previously [32, 35]. The PET tracer was injected intravenously as a bolus in an antecubital vein. There were no differences in the radioactivity between LLD (mean 149.5 MBq, SD = 4.7, range = 139–159) and controls (mean 150.6 MBq, SD = 5.1, range = 139–164) (p = 0.28). Image acquisition started 90 minutes after tracer injection and lasted for 30 minutes in list mode. Prior to the PET scan, a low-dose computed tomography scan was performed for attenuation correction. Random and scatter corrections were also applied. Images were reconstructed using Ordered Subsets Expectation Maximization (4 iterations X 16 subsets).

All PET frames were analyzed using an in-house Matlab® script (run on Matlab 2017b) and SPM12. All six PET frames were realigned to the first frame to correct for potential head motion, and then averaged to create a single PET image. Because both limited spatial resolution of the PET scanner and GM atrophy in our population may lead to a partial volume effect (PVE) [36], PET images were subsequently corrected for PVE using the modified method of Muller-Gartner et al. [37] as implemented in our in-house Matlab script. After coregistration of the individual PET images to their respective T1-weighted MRI images, PET images were normalized to Montreal Neurological Institute space by using the transformation parameters calculated during T1 normalization. SUVR images were calculated using cerebellar GM as reference region. The cerebellar GM volume of interest was derived from the Automated Anatomical Labeling (AAL) atlas and was masked with the normalized subject-specific segmented GM map. SUVR images were smoothed with an 8 mm FWHM Gaussian kernel.

### Statistical Analysis

Descriptive statistics were used to analyze the clinical data. Distributions of all variables were inspected using histograms, q-q plots, and Shapiro-Wilk tests. Whole-brain analyses were conducted using SPM12, and other analyses were conducted by using R version 3.4.3.

To investigate any differences in GMV and amyloid deposition between groups, whole-brain voxel-wise comparisons of GMV and [^18^F]-flutemetamol uptake between LLD and healthy controls were performed by using SPM12, with age as a covariate. TIV and scanner type were included as additional covariates in the GMV analysis. ApoE *ε*4 carrier status was included as an additional covariate in the amyloid analysis. The significance threshold for the voxel-wise whole-brain analyses was set at a cluster-level family-wise error (FWE)-corrected p <0.05 with an individual voxel threshold of p = 0.001. Only voxel values greater than 0.1 were analyzed using absolute threshold masking in the GMV analysis.

Mean voxel values within the clusters determined to be significantly different between groups were extracted and used in subsequent analyses. First, group comparisons of amyloid deposition in regions of significantly lower GMV in LLD were conducted by using a Mann-Whitney U test and an analysis of covariance (ANCOVA) controlling for age and ApoE *ε*4 carrier status. Bonferroni correction was conducted to control for multiple comparisons (alpha = 0.05/the number of regions with significantly lower GMV in LLD).

Second, to investigate the associations between regional GMV differences and clinical factors in LLD, a series of multivariate analysis of covariance (MANCOVA) was conducted. This included 1) episodic memory domain score, 2) GDS, 3) late onset (>55 years) or early onset of depression, and 4) the presence of psychosis as independent variables. Age and TIV were included as covariates. For these analyses, we set p <0.0125 (alpha = 0.05/4) as a statistically significance threshold. We also conducted the same sequential MANCOVAs including age and ApoE *ε*4 carrier status as covariates and using SUVR values from the regions of significantly lower GMV in LLD to investigate potential correlations between regional amyloid deposition and clinical factors.

Finally, to investigate the relationship between GMV and amyloid deposition in LLD, partial correlation analyses (Spearman) including age, TIV and ApoE *ε*4 carrier status as covariates, were conducted by using extracted voxel values from regions of significantly lower GMV in LLD identified in the whole-brain analysis. Bonferroni correction was conducted to control for multiple comparisons.

## Results

We included 48 patients with LLD (mean age 74.1, SD = 7.5; 33 females; 28 late-onset depression; 16 psychotic depression), and 52 healthy controls (mean age 72.4, SD = 6.4). The mean GDS score was 23.1 (SD = 4.3). LLD showed significant lower episodic memory scores than healthy controls (t = −6.4, p <0.001). Episodic memory scores did not correlate with GDS scores (Spearman’s ρ = −0.08, p = 0.63) in LLD. Clinical characteristics of the participants are presented in Table 1.

**Table 1.** Clinical characteristics of participants

|  | Patients with LLD | Healthy controls | p-value |
| --- | --- | --- | --- |
| Number | 48 | 52 |  |
| Age, years (mean, SD) | 74.1 (7.5) | 72.4 (6.4) | 0.21 |
| Female, n (%) | 33 (68.5%) | 37 (71.2%) | 0.97 |
| ApoE ε4 carrier, n (%) | 12 (25.0%) | 22 (42.3%) | 0.12 |
| Psychotic depression, n (%) | 16 (33.3%) |  |  |
| Late-onset depression, n (%) | 28 (58.3%) |  |  |
| Geriatric Depression Scale<br>(mean, SD) | 23.1 (4.3) | 3.0 (3.1) | <0.001 |
| Episodic memory domain scores<br>(mean, SD) | -3.8 (3.0) | 0 (2.7) | <0.001 |

Abbreviations: SD, standard deviation

### Regional differences in amyloid deposition and GMV

Whole-brain voxel-wise analysis revealed that patients with LLD showed significantly lower GMV in the left posterior temporal region (mainly ventral and lateral cortex), left parietal region (mainly at the temporo-parietal junction), and right ventral occipital region compared to healthy controls (Fig. 1A–E; Table 2). Brain regions showing higher GMV in LLD than healthy controls were not detected. These findings were minimally dependent on template, smoothing extent, scanner effect or amyloid positivity status (Supplementary information). In contrast, no statistically significant group differences in amyloid deposition were detected in the whole-brain analysis. Moreover, SUVR values in the regions of statistically significant lower GMV in LLD did not differ between groups (Fig. 1F–H; Supplementary information).

**Table 2.** Regions of significantly lower GMV in LLD detected in the whole-brain GMV analysis.

| Cluster Size<br>(voxels) | Brain Regions | Peak MNI coordinates |  |  | T values | Cluster-level p | Voxel-level p |
| --- | --- | --- | --- | --- | --- | --- | --- |
|  |  | x | y | z |  | (FWE-corrected) | (FWE-corrected) |
| 3295 | Left temporal cluster |  |  |  |  | <0.001 |  |
|  | Left middle temporal gyrus | -64 | -50 | 6 | 5.51 |  | 0.004 |
|  |  | -60 | -57 | -4 | 4.58 |  | 0.12 |
|  | Left inferior temporal gyrus | -50 | -62 | -8 | 4.81 |  | 0.055 |
|  |  | -38 | -46 | -14 | 4.14 |  | 0.40 |
|  |  | -51 | -58 | -22 | 3.96 |  | 0.57 |
|  |  | -50 | -51 | -8 | 3.77 |  | 0.77 |
|  |  | -46 | -50 | -27 | 3.39 |  | 0.98 |
|  | Left fusiform gyrus | -22 | -44 | -14 | 4.61 |  | 0.11 |
|  |  | -34 | -62 | -9 | 3.99 |  | 0.54 |
| 1399 | Left parietal cluster |  |  |  |  | 0.005 |  |
|  | Left middle temporal gyrus | -51 | -50 | 22 | 5.42 |  | 0.006 |
|  | Left supramarginal gyrus | -56 | -46 | 32 | 4.53 |  | 0.14 |
|  |  | -58 | -57 | 28 | 3.62 |  | 0.89 |
| 1564 | Right occipital cluster |  |  |  |  |  |  |
|  | Right lingual gyrus | 14 | -86 | -10 | 4.37 | 0.003 | 0.22 |
|  | Right middle occipital gyrus | 32 | -100 | -2 | 4.09 |  | 0.44 |
|  | Right inferior occipital gyrus | 24 | -93 | -6 | 3.99 |  | 0.54 |
|  |  | 30 | -86 | -10 | 3.79 |  | 0.75 |
|  |  | 44 | -75 | -14 | 3.77 |  | 0.77 |
|  | Right fusiform gyrus | 30 | -75 | -16 | 3.91 |  | 0.63 |
|  |  | 46 | -58 | -20 | 3.86 |  | 0.68 |

Peak coordinates were local maxima more than 10 mm apart.

**Fig. 1.**
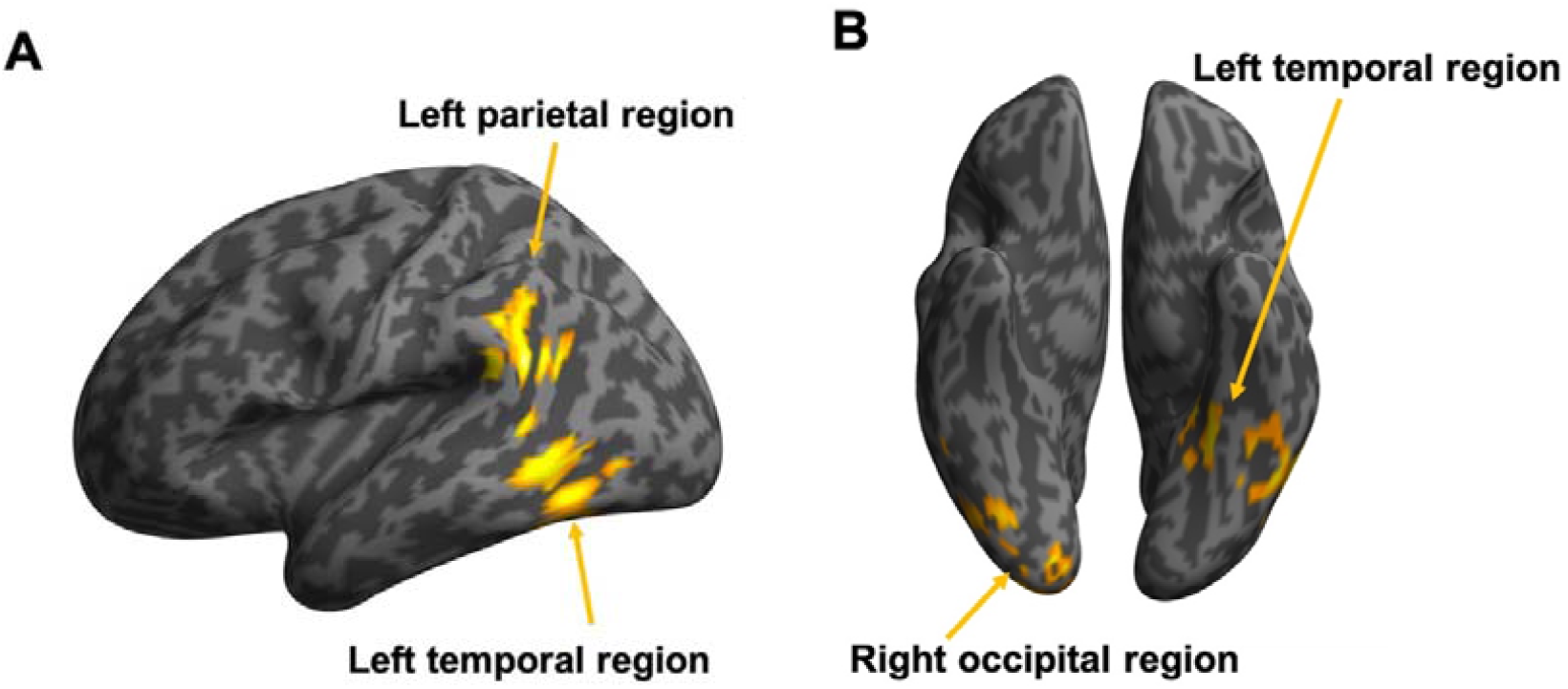
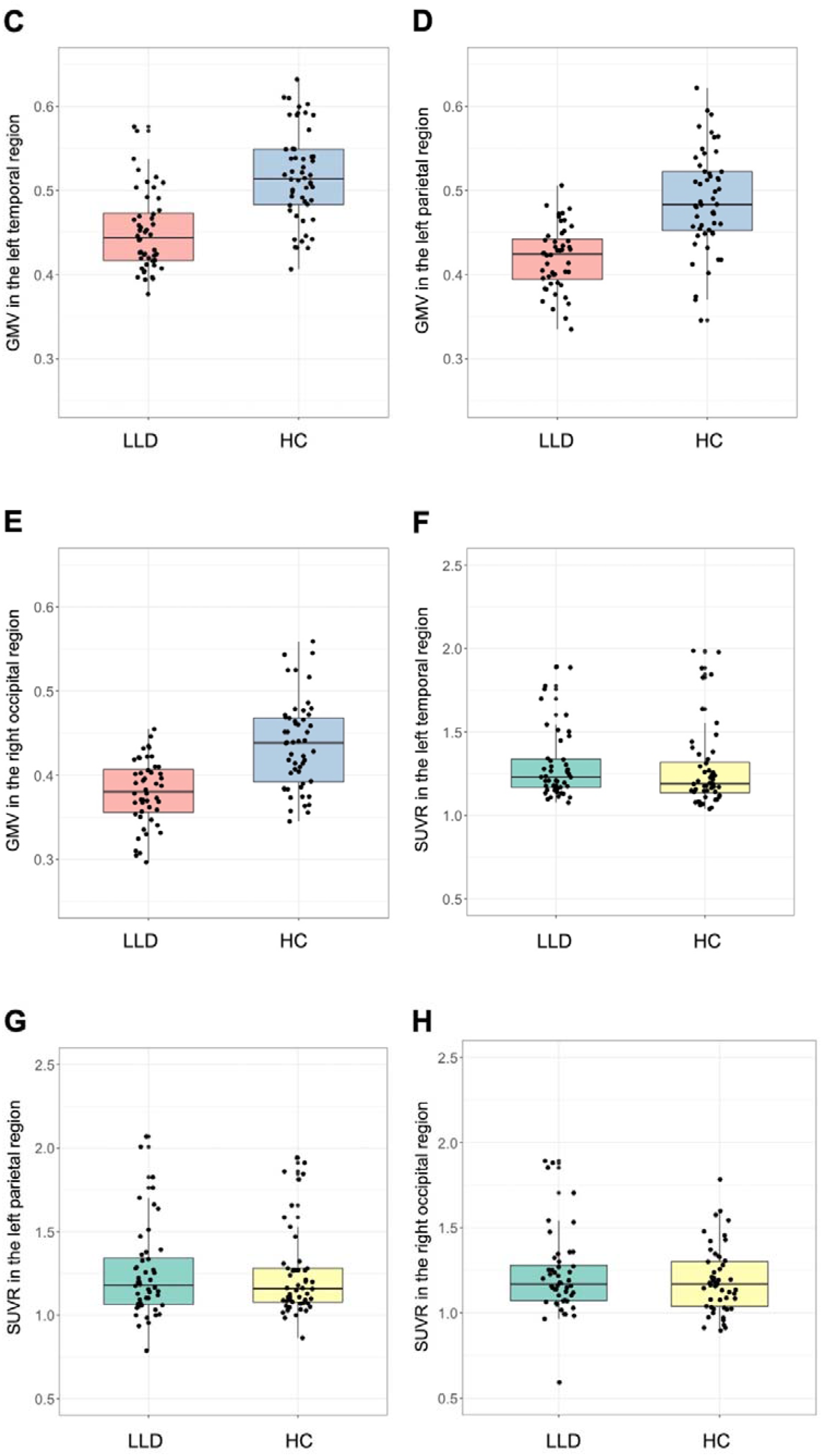
Brain regions showing significantly lower GMV in patients with late-life depression compared to healthy controls following whole-brain voxel-wise analysis. Three regions were identified: the left temporal, left parietal and right occipital regions. Areas associated with lower GMV in LLD are shown in yellow in (A) the left lateral view and (B) the bottom view. Boxplots of GMVs and SUVRs in each region are also presented: GMV in (C) the left temporal, (D) the left parietal, and (E) the right occipital regions; SUVR in (F) the left temporal region, (G) the left parietal region, and (H) the right occipital region. GMVs in these regions were significantly lower in LLD than in healthy controls (C–E), whereas SUVRs in these regions did not differ between groups (F–H). Y-axis represents mean voxel values in each cluster. Abbreviations: HC, healthy control; LLD, late-life depression

### Associations of clinical factors with GMV/amyloid deposition in LLD

There was a significant main effect of episodic memory scores on regional GMV differences (F_3, 34_ = 4.6, p = 0.008). Separate ANCOVAs revealed that episodic memory scores were significantly associated with GMV in the left temporal region (F_1, 36_ = 12.3, p = 0.001). The association between episodic memory and GMV did not survive after multiple comparison correction in the right occipital region (F_1, 36_ = 6.5, p = 0.015), and there was no association with GMV in the left parietal region (F_1, 36_= 0.96, p = 0.33). There were no main effects of GDS (F_3, 33_ = 0.15, p = 0.93), age at onset (F_3, 42_ = 0.10, p = 0.97), or the presence of psychosis (F_3, 42_ = 0.30, p = 0.82). To illustrate the results, scatter plots of clinical factors and GMV in the left temporal region are presented in Fig. 2. There were no significant main effects of any clinical factors on regional amyloid deposition (Supplementary information).

**Fig. 2.**
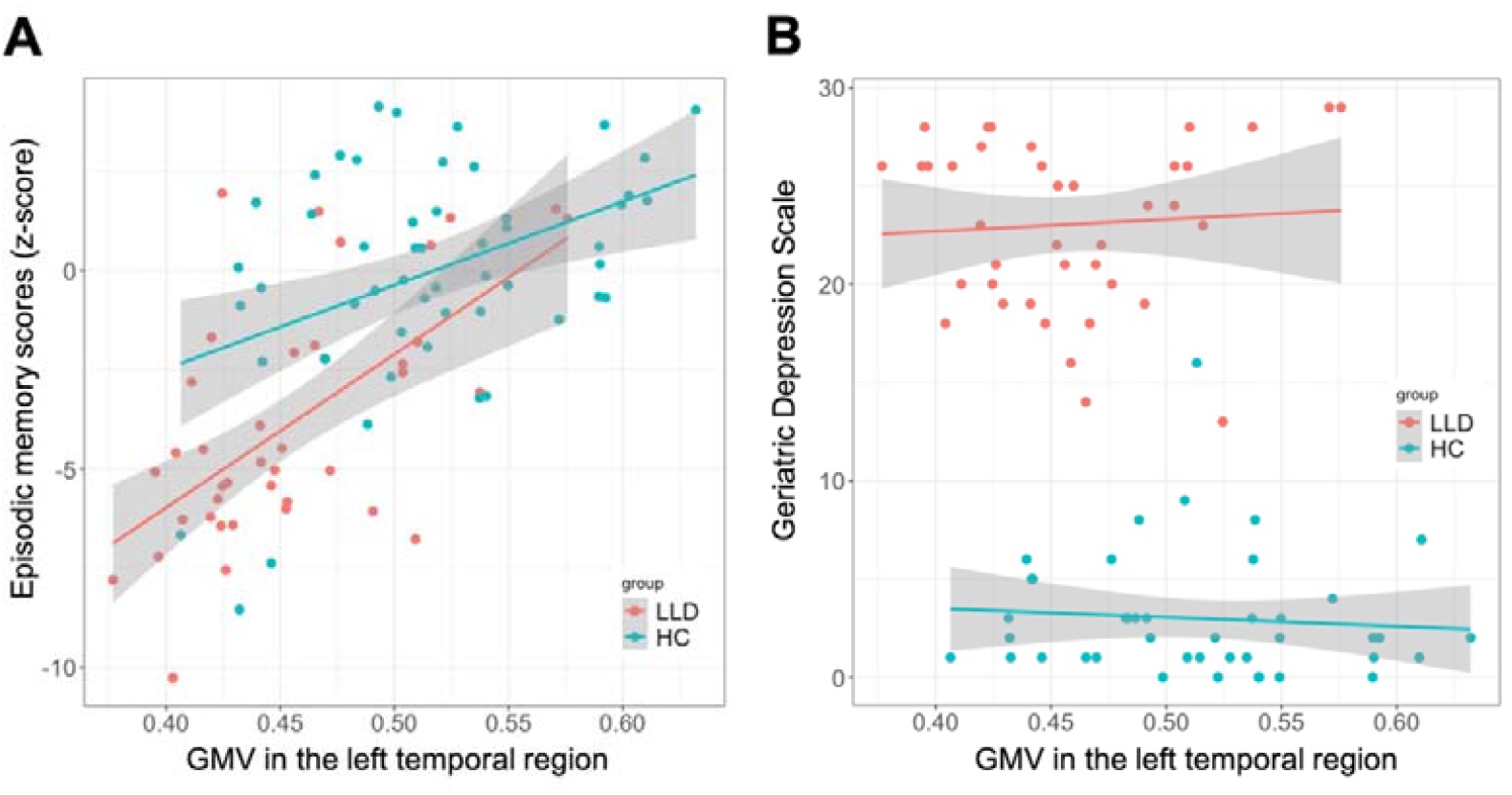
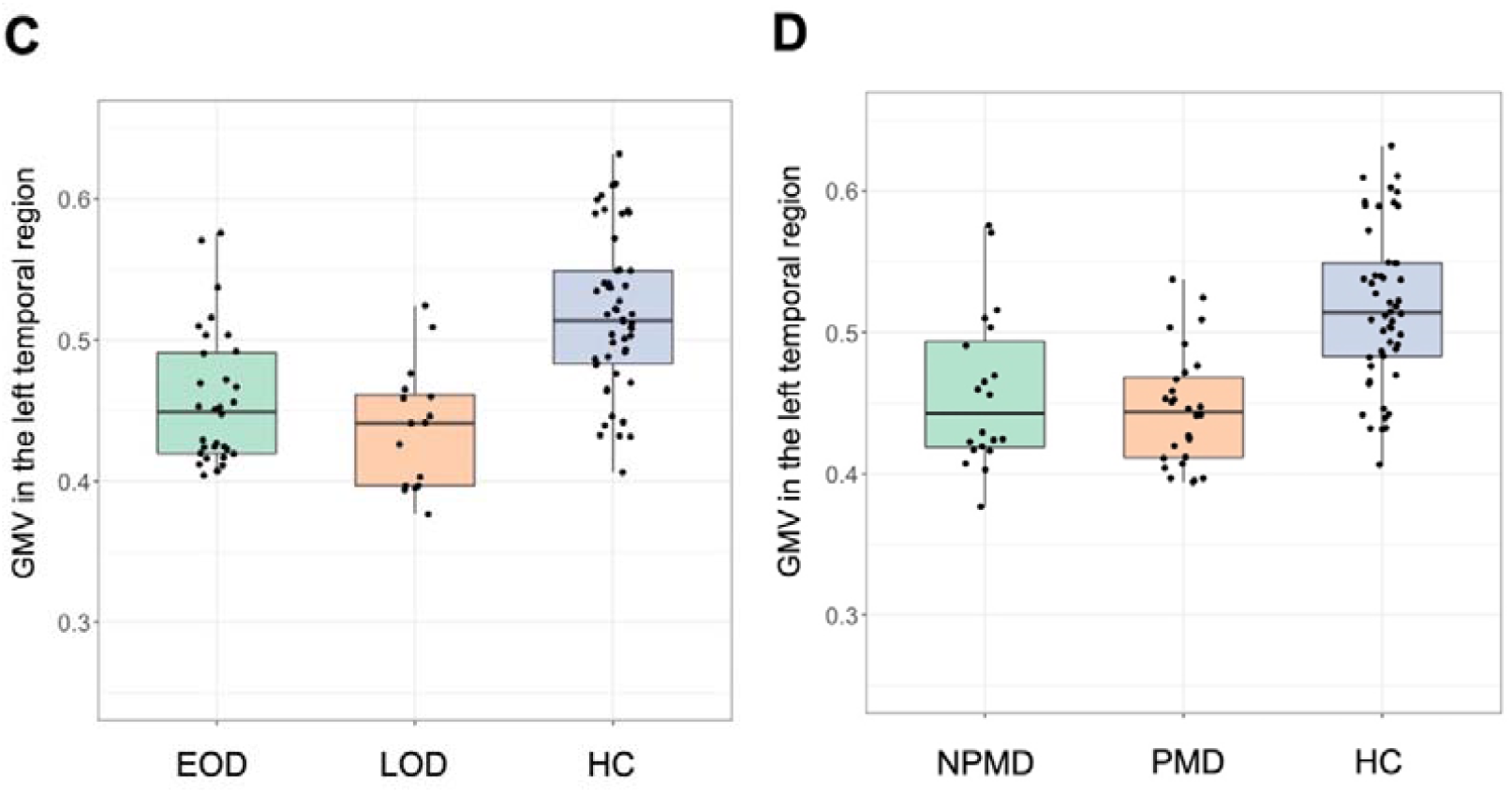
Associations of clinical factors with regional GMV decrease in the left temporal region. Episodic memory scores correlated with lower GMV in the left temporal region. To illustrate the results of the MANCOVAs, scatter plots of (A) episodic memory scores and (B) GDS, and boxplots of (C) onset age and (D) the presence of psychosis are presented. Episodic memory scores (A) had a significant main effect on regional GMV difference, whereas other clinical factors (B–D) did not have significant main effect. In all figures, data from healthy controls are also overlaid for reference. Y-axis in Figure 2C and 2D represents mean voxel values in the left temporal cluster. Abbreviations: EOD, early-onset depression; GDS, geriatric depression scale; GMV, gray matter volume; HC, healthy control; LLD, late-life depression; LOD, late-onset depression; NPMD, non-psychotic major depression; PMD, psychotic major depression

### Associations between GMV and amyloid deposition

Within the LLD cohort, there were no associations between GMV and SUVR in the left temporal (p = 0.91), left parietal (p = 0.09) and right occipital regions (p = 0.73) (Supplementary Fig. 1).

## Discussion

In this study we aimed to extend the literature on the relationship between LLD and AD-pathophysiology by applying multimodal whole-brain assessments comparing patients with LLD to healthy controls. We found that LLD was associated with lower regional GMV but not with differences in amyloid deposition compared to healthy controls. Additionally, lower left temporal lobe GMV was associated with episodic memory impairment in LLD.

### AD-pathophysiology in LLD

Amyloid deposition is considered to be one of the earliest neuropathological hallmarks in AD [38], along with tau deposition, which triggers downstream effects, such as gray matter atrophy and cognitive impairment. Although amyloid pathology is hypothesized to be also related to the neurobiology of LLD [39], we did not find higher amyloid deposition in LLD compared to healthy controls in the whole brain analysis. Surprisingly, recent work [19] reported lower amyloid deposition in LLD. The discrepancy could be due to considerable differences in methodology. For example, Mackin et al. [19] analyzed data from multiple sites (33 imaging sites with 9 different scanner types) which could introduce methodological variability that is less readily controlled compared to a monocentric study. Additionally, the severity of depressive symptoms between our cohorts was significantly different (mean GDS of 7.3 in their study vs. 23.1 in our study: mild vs. severe depressive symptoms), and there were important differences in the quantification methods of amyloid burden (different PET tracers and the use of partial volume correction in our data).

Another recent work, including 21 moderately depressed elders with limited or no prior antidepressant treatment history, reported increased amyloid deposition in the left parietal cortex in LLD [40] and no differences in GMV, which is in contrast with our findings. It is possible that these subjects represent a different clinical phenotype to ours given their lack of prior medication use and preserved cognitive functioning relative to controls. Whilst the inconsistent findings in group differences in amyloid burden in LLD challenges the notion that amyloid pathology plays a central role in the disorder, it does not necessarily exclude a neurobiological link between LLD and AD.

Although hippocampal volume reduction has been the main focus in LLD and AD literature, GMV reduction beyond hippocampus has also been reported [22, 24]. Our findings of GMV decrease in LLD are in line with other studies, which reported GMV reduction in the left temporoparietal region [23, 41] and occipital region [42]. Temporoparietal and to a lesser degree occipital atrophy are also reported in AD literature [20, 43–45]. Therefore, the current results of GMV reduction in LLD partly overlap with atrophic patterns in AD. In the current whole-brain analysis using the stringent cluster-level FWE correction, the left hippocampal volume reduction in LLD we reported in our previous ROI analysis [32] was not detected. However, a supplementary ROI analysis confirmed our previous finding (Supplementary information).

### The etiology of GMV reduction in LLD

Stress has long been considered to play a major role in depression [46, 47] and stress-related neurotoxic effects, such as elevated cortisol levels, excess oxidative stress, and reduced neurotrophic factors, could contribute to GMV reduction [48]. If stress-mediated effects on the brain were the main explanation of GMV reduction, one would expect that patients with EOD would show lower GMV than those with LOD [49]. However, we did not find any associations between regional GMV and age at onset of depression. Another possible explanation is that lower GMV reduction arises from accelerated aging [50, 51]. Notably, consistent with a previous study [23], we found a negative correlation between age and GMV in our healthy cohort, which was not present in LLD i.e. the GMV differences were most pronounced between the younger patients compared to the younger controls and these differences diminished with increasing age (Supplementary information).

Our findings of GMV reduction unrelated to amyloid pathology may also be related to a condition called suspected non-Alzheimer’s disease or non-amyloid pathophysiology (SNAP) [52, 53], which is related to an increased risk of cognitive decline and progression to dementia [53], and is estimated to affect almost a quarter of adults over 65 years of age [53]. Moreover, tau deposition in the MTL in the absence of cortical amyloid deposition was postulated as primary age-related tauopathy (PART) [54]. Tau deposition has been associated with temporal and parietal cortical atrophy in cognitively normal subjects [55] and left hippocampal atrophy in subjects with PART [56]. Depressive symptoms have also been related to tau accumulation in the entorhinal cortex and inferior temporal lobe in cognitively normal older adults using tau [^18^F] AV-1451 PET imaging [57]. A recent meta-analysis found limited evidence for tau pathology in LLD, but concluded it could not be ruled out given the lack of available data [58]. As others have hypothesized, it is plausible that a combination of neurobiological processes such as proteinopathies, vascular pathology and stress-mediated damage interact to lower the threshold for the development of neuropsychiatric symptoms in vulnerable individuals [59]. Conversely, in some individuals, reactive depressive symptoms may develop due to environmental stressors and contribute to transient cognitive dysfunction unrelated to neuropathology. Further research into non-amyloid pathology in LLD, and its association with stress, age at onset of depression, and GMV reduction may help to elucidate the underlying neurobiology of LLD.

### Episodic memory dysfunction in LLD

Episodic memory dysfunction is considered as the first clinical sign of AD [21], and it is also reported in LLD [2–4]. We found episodic memory dysfunction in LLD, and this was associated with lower GMV in the left temporal region, but not with amyloid burden. Our results are supported by previous studies, showing the associations of episodic memory dysfunction with reduced hippocampal or parahippocampal volume [34, 60]. The identified temporal region in the current study included lateral temporal regions as well as the parahippocampal gyrus. Interestingly, recent whole-brain functional imaging studies have revealed that large-scale distributed networks, including lateral temporal or temporoparietal regions, were associated with episodic memory [61–63]. Notably, the auditory verbal learning task used in our study involves various cognitive processes, including semantic processing and working memory [64]. Auditory verbal short-term memory has been associated with activations in left-lateralized posterior superior temporal regions and inferior parietal/supramarginal gyrus [65] as well as with left lateral temporal and temporoparietal regions in patients with AD [64].

We found that regional GMV differences in LLD correlated with impairments in episodic memory but not with depression severity, and that episodic memory scores did not correlate with depression severity. This is in contrast to a previous meta-analysis of depression at all ages which found an association between depression severity and cognitive function [66]. However, our negative finding is consistent with previous studies which focused specifically on LLD [2, 34], suggesting that the effect of depression severity on cognitive function may be age-dependent. Moreover, our results suggest that the neural correlates mediating affective symptoms and cognitive symptoms in LLD might be different.

### Strengths and limitations

It is important to emphasize the uniqueness of our cohort when comparing our analysis with previous studies. Our main focus was to explore the contribution of AD-associated pathophysiology to the neurobiology of LLD. Therefore, we recruited actively depressed patients without a diagnosis of MCI or dementia. In contrast, most previous amyloid-PET studies have focused on a history of depression or depressive symptoms in healthy subjects [13–15] or subjects who were already diagnosed with MCI or AD at the time of scanning [67, 68]. However, there is a possibility that the underlying neurobiology of a psychiatric disorder (e.g. LLD) with/without cognitive dysfunction and a neurodegenerative disorder (e.g. AD) with depressive symptoms are different. In this context, Mackin et al [19] and our previous study [32] have investigated LLD patients with mild or severe depressive symptoms respectively, and neither study found higher amyloid to be associated with LLD, whilst Smith et al [40] reported higher amyloid in LLD with moderate depressive symptoms without prior antidepressant treatments in a smaller cohort of 21 patients and 21 controls.

One potential limitation of our study is the cross-sectional design, therefore there is still a possibility that amyloid deposition is related to longitudinal GM atrophy, onset of depression, and cognitive dysfunction. Second, most participants took antidepressant medications, which might have an effect on amyloid accumulation [69], although no association between the length of lifetime antidepressant treatments and amyloid burden in LLD was reported recently [19]. Third, whilst we attempted to mitigate any bias due to including data from 19 control subjects scanned on a different scanner, in the absence of formal test data from a subject scanned on both scanners to confirm this, we cannot rule out such effects. Fourth, we did not find any differences in GMV or amyloid deposition between psychotic depression and non-psychotic depression, nor those between EOD and LOD. These negative findings might be due to the limited sample size in each group. In addition, the negative finding related to amyloid deposition might be a consequence of the heterogeneity of our cohort and therefore may not be generalized to all LLD as a whole. Future investigation of a more homogenous LLD sample could provide further insights into the role of amyloid in specific LLD subtypes.

In conclusion, LLD was associated with lower GMV in regions also implicated in AD. Left temporal GMV reduction was associated with episodic memory dysfunction in LLD. However, amyloid deposition did not differ between LLD and healthy subjects, and it did not correlate with clinical symptoms nor clinical characteristics of LLD. Our results suggest that amyloid deposition might not play a central or proximal role in the neurobiology of LLD.

## Data Availability

The datasets generated during and/or analyzed during the current study are not publicly available.

## Trial registration

European Union Drug Regulating Authorities Clinical Trials identifier: EudraCT 2009-018064-95.

## Data Availability

The datasets generated during and/or analyzed during the current study are not publicly available.

## Author Contributions

M. Vandenbulcke, L. Emsell, J. Van den Stock, FL. De Winter, F. Bouckaert, A. Takamiya and T. Vande Casteele designed the study. A. Takamiya, T. Vande Casteele, L. Emsell, and M. Koole analyzed the data. A. Takamiya and T Vande Casteele wrote the first draft. M. Koole, FL. De Winter, F. Bouckaert, J. Van den Stock, S. Sunaert, P. Dupont, R. Vandenberghe, K. Van Laere, M. Vandenbulcke, and L. Emsell contributed to the interpretation of the results and revised the first draft. All authors contributed to and have approved the final manuscript.

