## supplementary information for "Lower regional gray matter volume in the absence of higher cortical amyloid burden in late-life depression"

*Supplemental Information*

### Supplementary Results

#### Group differences in regional amyloid deposition in regions with lower gray matter in LLD

We extracted standardized uptake value ratio (SUVR) from three lower gray matter volume (GMV) regions identified in the whole-brain GMV analysis (See Table 1 and Figure 1 in the main text), and we compared them between patients with late-life depression (LLD) and healthy controls (HC). Mann-Whitney U-tests showed no significant differences in SUVRs in the left temporal region ($p = 0.18$), left parietal region ($p = 0.88$), and right occipital region ($p = 0.59$) between patients with LLD and HC (Figure 1 in the main text). An analysis of covariance (ANCOVA) controlling for age and ApoE $\varepsilon 4$ carrier status also did not show any differences in SUVRs in the left temporal region ($p = 0.60$), left parietal region ($p = 0.92$), and right occipital region ($p = 0.46$) between groups.

#### Associations of clinical factors with amyloid deposition in LLD

There were no main effects of the episodic memory scores ($F_{3, 34} = 1.2$, $p = 0.32$), GDS ($F_{3, 33} = 0.50$, $p = 0.68$), age at onset of depression ($F_{3, 41} = 1.5$, $p = 0.22$), or presence of psychosis ($F_{3, 41} = 2.6$, $p = 0.07$) on the regional amyloid deposition.

We also conducted additional whole-brain regression analyses to identify any potential associations between regional amyloid deposition and clinical scores (i.e., episodic memory scores and GDS), including age, ApoE $\varepsilon 4$ status, and diagnosis as covariates. In these analyses, we did not find any significant correlations between clinical scores and amyloid deposition with significance threshold at a cluster-level FWE-corrected $p < 0.05$ with an individual voxel threshold of $p = 0.001$.

**Associations between GMV and amyloid deposition**

There were no correlations between GMV and SUVRs in the left temporal region ($\rho = -0.02$, $p = 0.91$), left parietal region ($\rho = 0.26$, $p = 0.09$), and right occipital region ($\rho = 0.05$, $p = 0.73$) (Supplementary Figure S1).

**Supplementary Figure S1.** Associations between gray matter volume (GMV) and amyloid deposition in the lower GMV regions in patients with late-life depression compared with healthy controls. There were no correlations between GMV and SUVR in the (A) left temporal region; (B) left parietal region; and (C) right occipital region. Data from healthy controls are also overlaid as references.

GMV, gray matter volume; LLD, late-life depression; HC, healthy controls

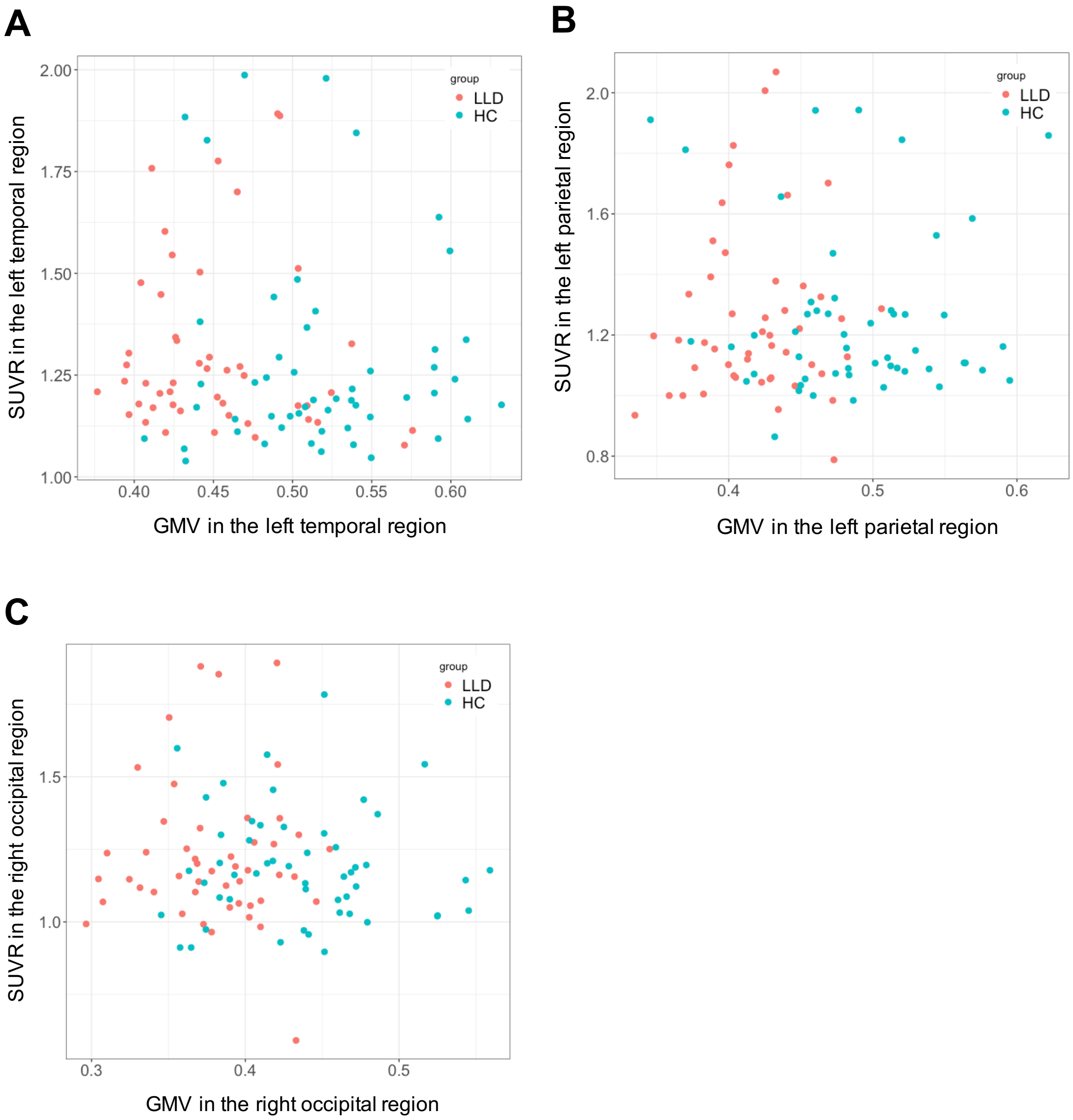


**Effects of smoothing parameters on whole-brain GMV and amyloid analyses**

To investigate the potential effects of smoothing parameters on the main results, we re-analyzed our data with different smoothing parameters: i.e. a Gaussian kernel of 6mm or 10mm full-width at half maximum (FWHM). In the whole-brain GMV analyses, the left temporoparietal region was detected with both smoothing parameters (Supplementary Table S1; Supplementary Table S2; Supplementary Figure S2; Supplementary Figure S3). The whole-brain analysis of GMV images with a 6 mm

FWHM Gaussian kernel, the left medial temporal region, right thalamus, and left cerebellum were also detected. Whole-brain group comparisons in amyloid deposition with any smoothing parameters did not show any differences between patients with LLD and healthy controls.

**Supplementary Table S1.** Results of whole-brain analysis using GMV images with a 10 mm FWHM Gaussian kernel.

| Cluster Size (voxels) | Brain Regions | Peak MNI coordinates x | y | z | T values | Cluster-level p (FWE-corrected) | Voxel-level p (FWE-corrected) |
|---|---|---|---|---|---|---|---|
| 7925 | Left temporoparietal cluster | | | | | <0.001 | |
| | Left middle temporal gyrus | –51 | –50 | 22 | 5.53 | | 0.002 |
| | | –66 | –50 | 6 | 5.37 | | 0.004 |
| | | –58 | –57 | –3 | 4.59 | | 0.068 |
| | | –66 | –28 | –3 | 3.46 | | 0.866 |
| | Left inferior temporal gyrus | –50 | –62 | –8 | 4.82 | | 0.032 |
| | | –51 | –60 | –21 | 4.12 | | 0.273 |
| | Left fusiform gyrus | –22 | –45 | –12 | 4.62 | | 0.062 |
| | | –36 | –50 | –12 | 4.18 | | 0.233 |
| | | –34 | –64 | –8 | 4.08 | | 0.298 |
| | Left superior temporal gyrus | –52 | –32 | 14 | 3.97 | | 0.394 |
| | Left Inferior parietal lobule | –56 | –36 | 40 | 3.55 | | 0.79 |
| 2604 | Right occipital cluster | | | | | 0.003 | |
| | Right inferior occipital lobe | 24 | –93 | –6 | 4.1 | | 0.287 |
| | | 42 | –75 | –15 | 3.8 | | 0.554 |
| | | 30 | –86 | –12 | 3.77 | | 0.58 |
| | Right lingual gyrus | 14 | –86 | –10 | 3.85 | | 0.5 |
| | Right fusiform gyrus | 46 | –58 | –20 | 3.7 | | 0.649 |
| | | 34 | –66 | –4 | 3.45 | | 0.872 |

**Supplementary Table S2.** Results of whole-brain analysis using GMV images with a 6 mm FWHM Gaussian kernel.

| Cluster Size (voxels) | Brain Regions | Peak MNI coordinates x | y | z | T values | Cluster-level p (FWE-corrected) | Voxel-level p (FWE-corrected) |
|---|---|---|---|---|---|---|---|
| 1335 | Left temporal cluster | | | | | <0.001 | |
| | Left middle temporal gyrus | –63 | –50 | 6 | 5.42 | | 0.012 |
| | | –48 | –69 | 0 | 4.5 | | 0.257 |
| | | –68 | –46 | –9 | 3.33 | | 1 |
| | Left inferior temporal gyrus | –50 | –62 | –8 | 4.79 | | 0.109 |
| | | –60 | –56 | –6 | 4.6 | | 0.195 |
| | | –52 | –58 | –22 | 3.85 | | 0.888 |
| | | –51 | –51 | –6 | 3.65 | | 0.977 |
| | Left fusiform gyrus | –40 | –64 | –14 | 3.54 | | 0.994 |
| 813 | Left temporoparietal cluster | | | | | 0.003 | |
| | Left middle temporal gyrus | –52 | –51 | 21 | 4.83 | | 0.096 |
| | Left supramarginal gyrus | –57 | –45 | 30 | 4.31 | | 0.428 |
| | | –57 | –57 | 30 | 3.69 | | 0.966 |
| | Left inferior parietal lobule | –52 | –50 | 39 | 3.93 | | 0.824 |
| | Left angular gyrus | –44 | –57 | 27 | 3.7 | | 0.966 |
| 450 | Left occipitotemporal cluster | | | | | 0.044 | |
| | Left fusiform gyrus | –22 | –42 | –15 | 4.5 | | 0.261 |
| | | –27 | –51 | –12 | 4.27 | | 0.468 |
| 501 | Cerebellum cluster | | | | | 0.029 | |
| | Left cerebellum | –36 | –68 | –60 | 4.43 | | 0.316 |
| | | –46 | –69 | –46 | 3.38 | | 1 |
| | | –21 | –64 | –56 | 3.31 | | 1 |
| 531 | Left occipitotemporal cluster | | | | | 0.023 | |
| | Left fusiform gyrus | –34 | –10 | –33 | 4.33 | | 0.406 |
| | | –27 | –4 | –40 | 3.73 | | 0.953 |
| | Left inferior temporal gyrus | –33 | 3 | –36 | 3.59 | | 0.989 |
| 658 | Thalamic cluster | | | | | 0.009 | |
| | Right thalamus (VPL) | 15 | –18 | –2 | 4.17 | | 0.571 |
| | Right thalamus (MD) | 3 | –18 | 6 | 3.7 | | 0.964 |
| | Right thalamus (Pu) | 10 | –28 | 6 | 3.36 | | 1 |

VPL, ventral posterolateral; MD, mediodorsal nucleus; Pu, pulvinar nucleus

**Supplementary Figure S2.** Results of whole-brain analysis using GMV images with a 10 mm FWHM Gaussian kernel. With this larger smoothing kernel, left temporoparietal region was identified as one big cluster.

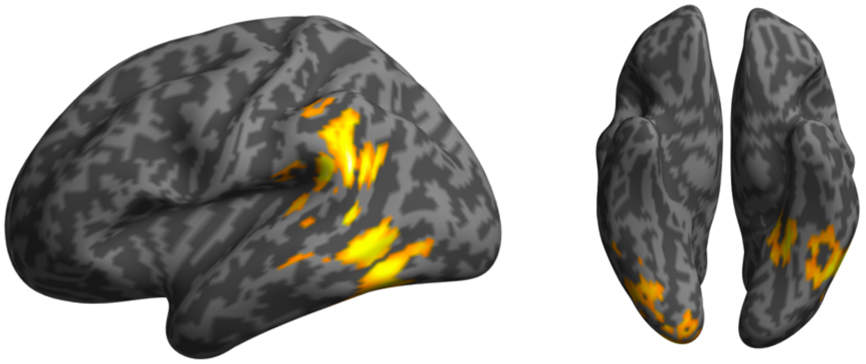

**Supplementary Figure S3.** Results of whole-brain analysis using GMV images with a 6 mm FWHM Gaussian kernel. With this smaller smoothing kernel, the left medial temporal region, right thalamus, and left cerebellum were also identified as lower GMV regions in LLD. The right thalamus and left cerebellum were not shown in the figure.

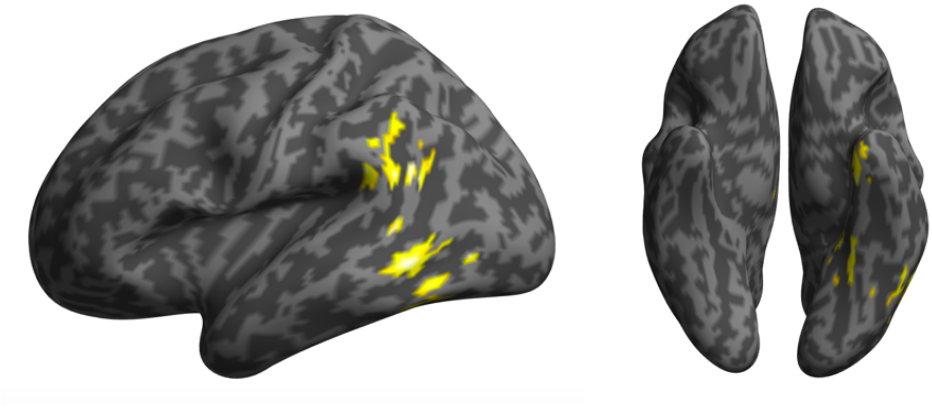

### Effect of dartel template on the results of GMV analyses

In the main analysis, we created a study-specific dartel template from all participants' data in this study because our cohort included only elderly subjects making the CAT12 default template based on healthy young volunteers suboptimal for our specific population. To investigate the potential effect of the dartel template in our GMV analysis, we conducted a whole-brain GMV analysis by using a default dartel template which is already implemented in CAT12. Even in this analysis, the left temporal and parietal regions were detected as lower GMV regions in patients with LLD (Supplementary Table 3; Supplementary Figure 4).

**Supplementary Table S3**. Results of whole-brain GMV analysis using default dartel template in CAT12.

| Cluster Size (voxels) | Brain Regions | Peak MNI coordinates x | y | z | T values | Cluster-level p (FWE-corrected) | Voxel-level p (FWE-corrected) |
|---|---|---|---|---|---|---|---|
| 2942 | Left temporal cluster | | | | | <0.001 | |
| | Left inferior occipital lobe | –46 | –74 | –4 | 4.86 | | 0.044 |
| | Left middle temporal gyrus | –64 | –48 | 6 | 4.51 | | 0.139 |
| | Left lingual gyrus | –22 | –48 | –10 | 4.43 | | 0.178 |
| | Left inferior temporal gyrus | –51 | –57 | –8 | 4 | | 0.522 |
| | | –48 | –48 | –28 | 3.93 | | 0.599 |
| | | –51 | –57 | –22 | 3.82 | | 0.709 |
| | Left middle temporal gyrus | –62 | –50 | –10 | 3.65 | | 0.864 |
| | Left fusiform gyrus | –36 | –45 | –14 | 3.53 | | 0.935 |
| 975 | Left parietal cluster | | | | | 0.003 | |
| | Left supramarginal gyrus | –54 | –45 | 24 | 4.32 | | 0.244 |
| | Left superior temporal gyrus | –57 | –32 | 16 | 3.99 | | 0.531 |
| | Left angular gyrus | –52 | –56 | 26 | 3.98 | | 0.542 |

**Supplementary Figure S4.** Results of whole-brain GMV analysis using the default dartel template in CAT12.

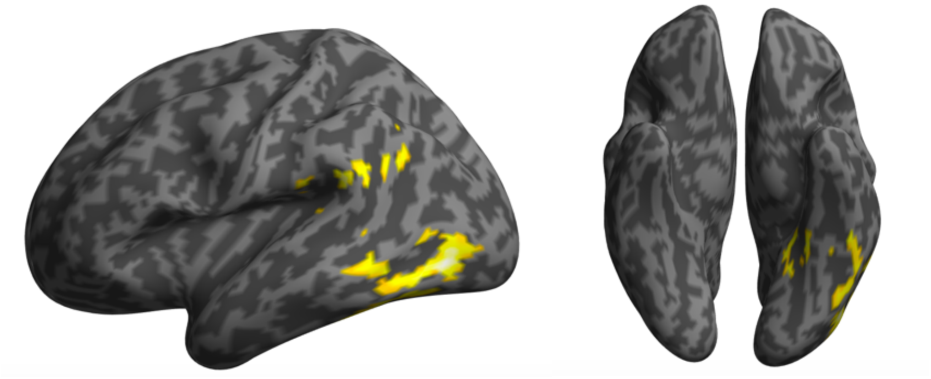

**Effects of different scanners on the results of GMV analysis**

In the main analyses, we accounted for the potential MRI scanner effect on the GMV analysis by including scanner as a covariate in the statistical model. We also conducted a whole-brain GMV analysis in a subpopulation excluding 19 subjects who received a different scanner with identical sequence parameters. Although the left temporal and parietal regions were not separated, the results were similar to the main results (Supplementary Table S4; Supplementary Figure 5).

**Supplementary Table S4.** Results of whole-brain GMV analysis excluding subjects who received a different scanner with identical sequence parameters.

| Cluster Size (voxels) | Brain Regions | Peak MNI coordinates x | y | z | T values | Cluster-level p (FWE-corrected) | Voxel-level p (FWE-corrected) |
|---|---|---|---|---|---|---|---|
| 5526 | Left temporoparietal cluster | | | | | <0.001 | |
| | Left middle temporal gyrus | –51 | –50 | 22 | 5.55 | | 0.006 |
| | | –63 | –50 | 6 | 5.44 | | 0.008 |
| | | –64 | –33 | –2 | 3.83 | | 0.753 |
| | Left inferior temporal gyrus | –60 | –56 | –6 | 4.71 | | 0.098 |
| | | –50 | –62 | –8 | 4.69 | | 0.102 |
| | | –50 | –51 | –8 | 4.22 | | 0.374 |
| | | –38 | –46 | –14 | 4.18 | | 0.405 |
| | | –51 | –58 | –22 | 3.97 | | 0.612 |
| | | –46 | –50 | –27 | 3.54 | | 0.951 |
| | Left superior temporal gyrus | –50 | –30 | 12 | 4.1 | | 0.489 |
| | Left fusiform gyrus | –22 | –45 | –12 | 4.67 | | 0.11 |
| | | –34 | –62 | –9 | 4.14 | | 0.449 |
| | Left supramarginal gyrus | –56 | –46 | 32 | 4.85 | | 0.063 |
| | | –58 | –57 | 30 | 3.48 | | 0.969 |
| | Left angular gyrus | –42 | –57 | 27 | 3.88 | | 0.712 |
| 1847 | Right occipital cluster | | | | | 0.001 | |
| | Right lingual gyrus | 14 | –86 | –10 | 4.42 | | 0.226 |
| | Right inferior occipital lobe | 24 | –93 | –6 | 4.22 | | 0.373 |
| | | 44 | –76 | –14 | 3.87 | | 0.714 |
| | | 30 | –86 | –10 | 3.68 | | 0.879 |
| | Right middle occipital lobe | 32 | –100 | –3 | 4.1 | | 0.482 |
| | Right fusiform gyrus | 46 | –58 | –20 | 4.31 | | 0.303 |
| | | 30 | –75 | –16 | 3.91 | | 0.674 |

**Supplementary Figure S5.** Results of whole-brain GMV analysis excluding subjects who received a different scanner with identical sequence parameters.

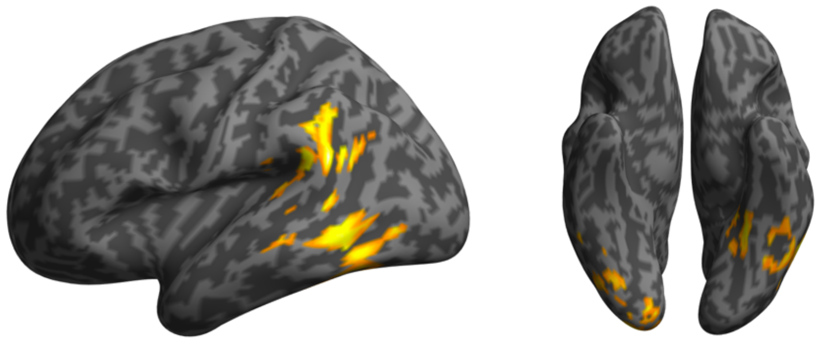

**Regional GMV difference in a subgroup excluding amyloid positive subjects**

Although there were no significant differences in the proportion of amyloid positive subjects between groups, we conducted the whole-brain GMV analysis in a subpopulation excluding 10 LLD subjects with amyloid positive and 12 healthy controls with amyloid positive. In addition to the left temporal, left parietal, and right occipital region, the right thalamus and left cerebellum were also detected as lower GMV regions in patients with LLD (Supplementary Table S5; Supplementary Figure 6).

**Supplementary Table S5.** Results of whole-brain GMV analysis excluding amyloid positive subjects.

| Cluster Size (voxels) | Brain Regions | Peak MNI coordinates x | y | z | T values | Cluster-level p (FWE-corrected) | Voxel-level p (FWE-corrected) |
|---|---|---|---|---|---|---|---|
| 1191 | Left parietal cluster | | | | | 0.009 | |
| | Left supramarginal gyrus | –51 | –50 | 22 | 5.41 | | 0.011 |
| | | –56 | –48 | 32 | 4.76 | | 0.088 |
| 2354 | Right occipital cluster | | | | | | |
| | Right lingual gyrus | 14 | –86 | –12 | 5.06 | <0.001 | 0.034 |
| | Right inferior occipital lobe | 30 | –86 | –10 | 4.38 | | 0.259 |
| | | 22 | –93 | –6 | 4.19 | | 0.413 |
| | | 42 | –66 | –16 | 3.89 | | 0.708 |
| | Right middle occipital lobe | 32 | –100 | –2 | 4.22 | | 0.382 |
| | | 39 | –93 | –2 | 3.71 | | 0.862 |
| | Right calcarine fissure and surrounding cortex | 8 | –90 | 0 | 3.81 | | 0.78 |
| | | 14 | –80 | 2 | 3.54 | | 0.952 |
| | | 16 | –104 | –2 | 3.46 | | 0.976 |
| | | 12 | –72 | 9 | 3.32 | | 0.994 |
| 1562 | Thalamic cluster | | | | | 0.002 | |
| | Right thalamus (VPL) | 15 | –18 | –2 | 4.95 | | 0.049 |
| 910 | Left temporal cluster | | | | | 0.026 | |
| | left inferior temporal gyrus | –51 | –62 | –8 | 4.34 | | 0.287 |
| | | –51 | –62 | –22 | 4.04 | | 0.562 |
| 959 | Cerebellum cluster | | | | | 0.022 | |
| | Left cerebellum | –28 | –57 | –46 | 4.24 | | 0.366 |
| | | –36 | –70 | –58 | 4.03 | | 0.568 |
| | | –18 | –54 | –50 | 3.36 | | 0.991 |

VPL, ventral posterolateral

**Supplementary Figure S6.** Results of whole-brain GMV analysis excluding amyloid positive subjects.

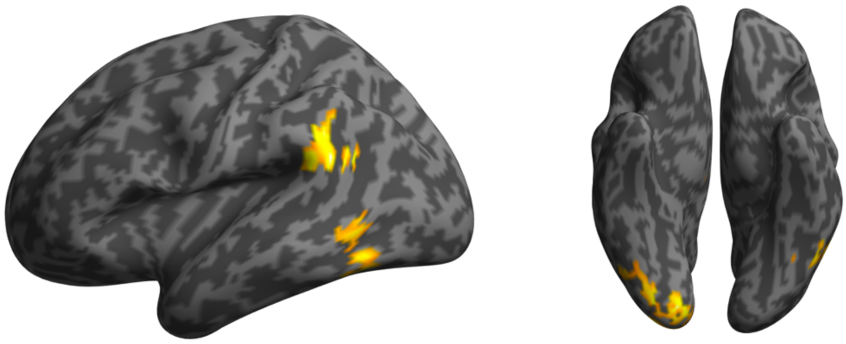

**Region of interest (ROI) analysis**

Since we reported hippocampal volume reduction in LLD in a previous paper (1), we also conducted a ROI analysis to confirm the previous finding by using another method (manual tracing vs. atlas-based automated calculation). In the current study, hippocampal volume was calculated based on automated anatomical labeling (AAL) atlas. Left hippocampal GMV was significantly lower in LLD compared to healthy controls, even after controlling for TIV and scanner type ($p = 0.018$). Right hippocampal GMV did not differ between groups ($p = 0.06$). Although the whole-brain analysis with a stringent statistical criteria did not detect hippocampal volume reduction in our main analysis, *a priori* ROI analysis with a different analysis method also detected left hippocampal volume reduction in LLD similar to our previous paper.

**Effect of age on GMV**

In general, age shows a negative correlation with GMV and cortical thickness (2–4). However, a previous study reported that there was no correlation between age and cortical thickness in patients with late-life depression (5). In the current study, as an additional analysis, we also found significant age-by-group interaction effects in the left temporal GMV ($p = 0.0058$), the left parietal GMV ($p < 0.001$), and the right occipital GMV ($p = 0.002$) (Figure S5A–C). Although this is based on cross-sectional data, our results might indicate an accelerating brain aging in patients with LLD (e.g., one potential interpretation is that patients with LLD in their 60s had similar GMV to healthy subjects in their 80s).

Figure S5. Differences in slopes of the age vs gray matter volume correlation between LLD and healthy controls. Additional analyses revealed significant age-by-group interaction effects in (A) the left temporal region, (B) the left parietal region, and (C) the right occipital region. Red lines represent LLD and turquoise lines represent healthy controls.

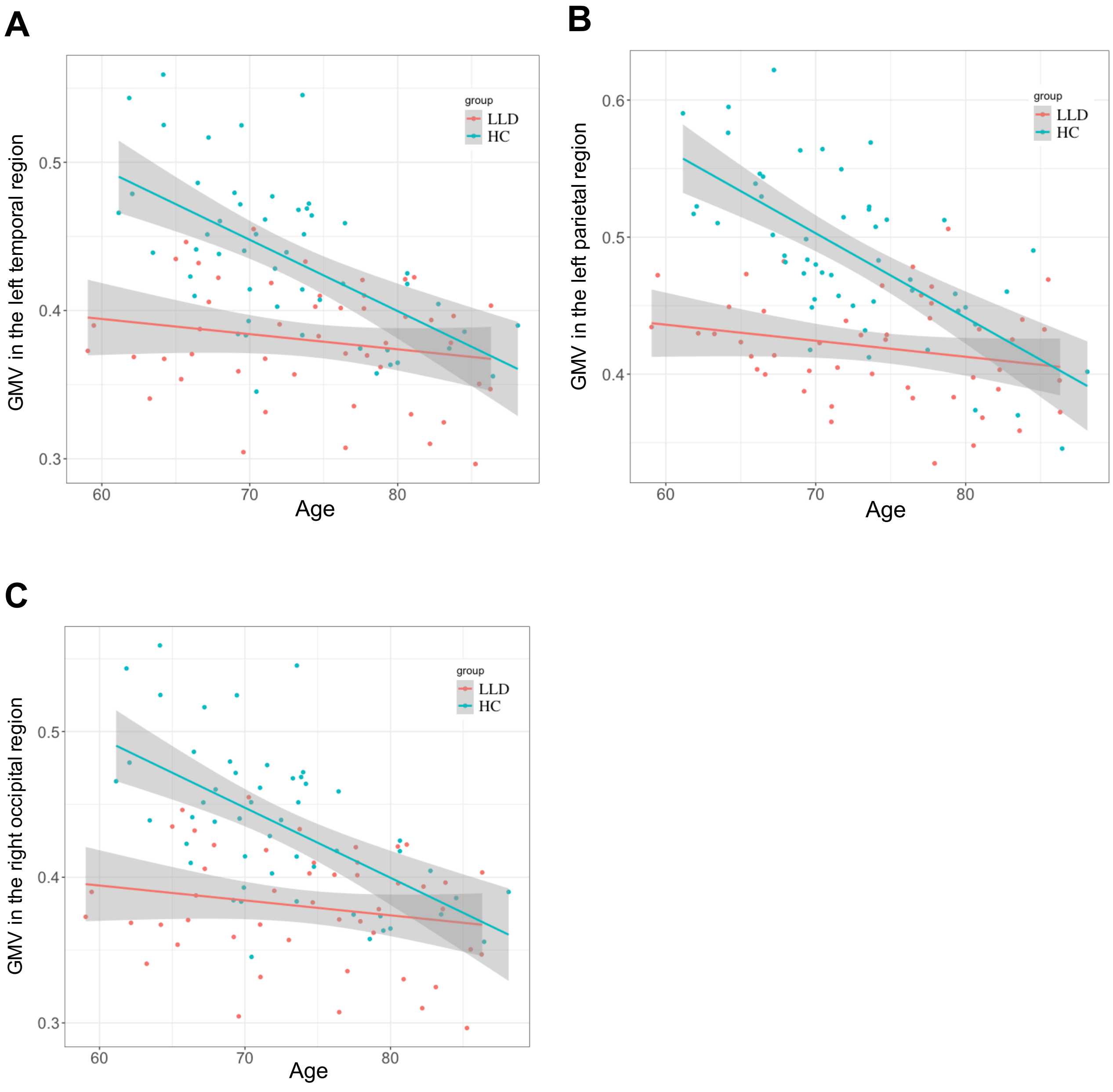
